# Measuring the Meaning of Genomic Results: Harmonization of the Metric for Case-Level Results in the CSER2 Consortium

**DOI:** 10.64898/2026.05.28.26354388

**Authors:** Bradford C. Powell, Laura M. Amendola, Katherine E. Bonini, David Crosslin, Lauren Desrosiers-Battu, Susan M. Hiatt, Lucia Hindorff, Eimear E. Kenny, Yusuph Mavura, Kathleen D. Muenzen Ferar, Neil Risch, Tamara Roman, Anne Slavotinek, Jessica Van Ziffle, Kevin M. Bowling

## Abstract

Yield of reported results from genetic testing provides a proximal measure of clinical usefulness. While ACMG/AMP guidelines provide representations of uncertainty for individual genetic variant classification, additional factors are considered when determining whether results explain a patient’s presentation. To standardize cross-consortium analysis, a working group of the Clinical Sequencing Evidence-Generating Research (CSER2) consortium iteratively identified factors used when contextualizing variant-level results to case-level interpretation (i.e., interpretation of an individual’s genetic data with respect to the indication for testing). Sites independently categorized results; complex cases were discussed collaboratively, leading to revision of classification categories. Our metric incorporates factors beyond classification of reported variants. Analogous to variant-level results, “Definitive Positive” and “Probable Positive” represent certainty that results may be clinically explanatory. The category “Inconclusive” applies when results may or may not fully explain the patient presentation, with subdivision into multiple (non-exclusive) subcategories. Cases falling outside all of the other categories are considered “Negative”. The overall diagnostic yield by this metric and use of categories for inconclusive results varied by CSER project, in part paralleling study design differences. This case-level categorization provides a meaningful assessment of diagnostic yield, and for inconclusive cases identifies potentially resolvable factors for case resolution.

## Introduction

The proximal goal of clinical genetic testing is to identify genetic variants that may be the underlying molecular cause of signs and symptoms that manifest in patients. While the ultimate goal of any medical intervention is to provide clinical utility, the diagnostic yield of genetic testing is a measure that can be used early in the process of assessing utility to establish an upper-limit for detecting individuals who could benefit from downstream interventions. It is therefore important to measure diagnostic yield accurately to determine the clinical utility or eventual societal benefit of genetic testing.

Guidelines for variant classification were published by the American College of Genetics and Genomics and the Association for Molecular Pathology (ACMG/AMP)^1^, and this formal structure for variant classification, as it has been adopted by clinical and research labs, has improved the consistency of pathogenicity classifications at the level of individual variants.^2,3^ However, the determination of whether a combination of genetic results (which can include panel sequencing, genome-scale sequencing, and microarray) is considered explanatory of some or all of the clinical features that provided the indication for testing depends on additional factors beyond variant pathogenicity, including confidence about the gene-disease association,^4^ expected mode of inheritance, disease-phenotype association, and, in the case of recessive conditions, the overall genotype, allelic state, and phase of genetic variants.^5^

The Clinical Sequencing Evidence-Generating Research consortium (CSER2) was a multi-institutional program developed to advance the science of translational genomics.^6^ The program’s focus represented an evolution from that of the earlier Clinical Sequencing Exploratory Research consortium^7^ in moving to address higher-level and downstream measures such as clinical and societal utility as genetic sequencing becomes more commonly used in clinical care. The consortium was funded by the National Human Genome Research Institute (NHGRI) in collaboration with the National Cancer Institute and the National Institute on Minority Health and Health Disparities, and was composed of six extramural and one NHGRI intramural research site along with a data coordinating center^8^, and affiliate members. Each of the research sites had individual research aims and heterogeneous study populations, but shared research questions required the standardization of a subset of measures. While other working groups within the consortium established harmonized measures of factors such as access to care, health literacy, information seeking, and quality of life^9^, the Sequence Analysis and Diagnostic Yield (SADY) working group was tasked with developing a shared representation of the diagnostic yield of testing performed at different sites to help characterize differences in yield that may result from a variety of covariates such as sequencing modality, ancestral genetic background of individuals being sequenced, proband vs. multisample testing strategy or different clinical indications for testing.

There are many instances in which genetic testing may identify combinations of variants that are not considered explanatory but may be “reportable” to the ordering provider and then to patients. These results are considered possibly explanatory or contributory, and returning these results to the ordering provider may serve to notify them that the results may be reclassified in the future^10^, or may prompt additional genetic studies or “reverse phenotyping^11^” to clarify the final results.

The development of a harmonized diagnostic yield metric served multiple purposes within the consortium. We found it critical to move beyond measures involving just the variants to the “case” as presented to the laboratories and clinicians: the interpretation of a research participant’s genetic data with respect to the indication for testing. Our metric was intended to improve consistency of reporting within the consortium to enable aggregation and comparison of data among sites. In addition, the shared terminology facilitated collaboration as we were able to discuss cases among sites more effectively. Furthermore, this metric was developed to better characterize how the different factors that can affect uncertainty about a case-level result can be communicated among testing laboratories, clinicians, and patients, and how these factors and classifications may change over time.

## Materials and Methods

The CSER2 SADY working group included members across all of the consortium sites, the data coordinating center, and affiliate members with research interests aligned to those of the CSER2 consortium. Working group members had expertise in clinical genetics, laboratory genetics and/or bioinformatics. The group reported to the CSER2 Steering Committee which included principal investigators from each of the sites as well as representatives from the National Institutes of Health. The categories and descriptions of this classification were developed through an iterative consensus process on monthly conference calls including representatives from all consortium sites, with primary development and refinement during 2018-2020. Proposed classifications were discussed in light of specific cases to help determine the clarity of category descriptions. Additional input on the classification was obtained through discussion with the CSER2 steering committee and by presentation at national professional society meetings.

Each site’s protocols were reviewed and monitored by its respective Institutional Review Board, and informed consent was obtained for human subjects participation and sharing of deidentified data. Because some aspects of research protocols at the individual sites were established prior to harmonization of the diagnostic yield measure, some specific aspects of scoring cases (e.g., who performs the classification, and at what time) were developed at individual sites (Table 1). Sites were encouraged to bring cases which they found difficult to classify to the SADY working group for discussion. Sites provided information on each participant’s phenotypic indication(s) for enrollment, de-identified demographic information, and case-level classifications as part of their quarterly progress reports. Sites that performed reanalysis or reclassification over the course of their study reported these updated results with standardized reasons for reclassification. This CSER2 standardized dataset was aggregated by the consortium data coordinating center and is available through the NHGRI Analysis Visualization and Informatics Lab-space (AnVIL; https://anvilproject.org/consortia/cser/resources). For the results presented in this manuscript, we relied only on this harmonized, deidentified dataset.

**Table 1.** Study design and protocols for classifying and updating case-level results classifications.

| CSER project | Study design summary | Procedures and personnel assigning phenotypic category | Study personnel making case-level classification | Time point(s) at which classification is made | Protocols for updating classifications |
| --- | --- | --- | --- | --- | --- |
| CHARM<sup>12</sup> | Germline ES vs. usual care evaluating for adults at risk for hereditary cancer | Not applicable (all participants were assigned category of "hereditary cancer risk assessment") | Study Genetic Counselor | Submission of quarterly progress reports | Classification may change based on variant reclassification as part of laboratory reanalysis protocols |
| KidsCanSeq<sup>13</sup> | Germline ES, tumor ES, microarray and transcriptome vs. targeted germline and/or | Clinical features abstracted from the electronic medical record (EMR). | Study genetic counselor and/or geneticist | Submission of quarterly progress reports | Study clinicians or GCs may prompt re-evaluation with additional clinical |
|  | tumor panel in children with cancer | Participants assigned to "Cancer" phenotype category (PC) plus additional PCs based on clinical features. |  |  | information. |
| NCGENES2<sup>14,15</sup> | Germline ES vs usual care in children with suspected genetic condition at first outpatient evaluation by pediatric genetics or neurology | Selected from available categories by enrolling clinician at initial visit | Clinical geneticist or molecular pathologist (in consultation with multidisciplinary sign-out committee and based on clinical laboratory report) | Prior to result disclosure to participants.. Re-assessed after discussion with study clinician and/or after secondary testing performed (e.g., parental testing to determine phase) | Study clinicians may prompt re-evaluation with additional clinical information. Sign-out committee meets with study clinicians on semi-annual basis to review results and case-level classification |
| NYCKidSeq<sup>16,17</sup> | Germline GS vs. targeted panel in children with suspected neurologic, immunologic and cardiac genetic conditions | Referring clinicians completed a phenotype checklist documenting the HPO terms relevant and provided it to the study team. Study genetic counselors collected a detailed 3-generation pedigree and additional clinical features at a pre-test counseling visit. Phenotype checklist, pedigree, and most recent referring clinician chart note sent to clinical labs to assist in variant interpretation. | Study genetic counselors. Study GCs could consult an interpretation committee of study physicians in medical genetics and/or pediatrics. | Prior to results disclosure to participants. | Should the clinical lab re-issue an amended report, the study genetic counselors re-evaluate SADY case classification and, if needed, consult with the study interpretation committee. |
| P<sup>3</sup>EGS<sup>18,19</sup> | Germline ES, singleton, duos or trios in infants and children with | Reviewed from referral forms; verified at the time of | Geneticist, genetic counselors, and laboratory | Prior to initial result reporting. Re-assessed after discussion | Study clinicians may request re-evaluation with additional |
|  | severe developmental disorders or in pregnancies in which the fetus has a structural anomaly | enrollment. HPO terms added by genetic counselor/study staff after consent | specialists (in consultation with multidisciplinary sign-out committee) | with study clinician and/or after secondary testing performed (e.g., parental testing to determine phase). | clinical information. |
| SouthSeq<sup>20</sup> | Germline GS in newborns with suspected genetic conditions | Selected from available categories (with an option to add additional HPO terms) by research nurse coordinator following enrollment and during chart review. | Variant Analysis Team | Prior to initial result reporting. | Study clinicians or GCs may prompt re-evaluation with additional clinical information. |

Differences in case-level classifications were visualized using UpSet^21^ plots with the Python UpSetPlot^22^ library version 0.9.0.

## Results

### Definition of categories in the case-level results metric

We chose terminology to distinguish between results at the variant level (described using the ACMG/AMP guidelines for classification of genetic variants) compared to case level results. While variant-level classification includes the terminology “Likely” to indicate some degree of uncertainty for individual variant pathogenicity, we chose the modifier “Probable” for situations which would typically be considered positive for the purposes of clinical management, but for which the classification is not considered absolute (e.g., there may still be some limited uncertainty at the variant level or with regard to complete phasing). Also, we chose “Inconclusive” to represent ambiguity at the case-level to avoid confusion with uncertainty of variant-level classification.

Results at the case-level were divided into four overall categories (Figure 1) based on whether the combination of identified genetic variants was considered to provide an explanation for the phenotype(s) present within the individual tested. The first two categories, “Definitive Positive” and “Probable Positive” would both typically be considered positive from the standpoint of enacting further medical management for a specific condition. In this manner, these categories are case-level analogs to the “Pathogenic” and “Likely Pathogenic” categories of ACMG/AMP variant-level classifications. Cases in which there is substantial ambiguity in the relationship between the genetic variants and patient’s phenotype are given the top-level classification of “Inconclusive”, with non-exclusive subcategories applied as reasons for ambiguity. Cases in which there are no identified variants with an expected relationship to the patient phenotype or reason(s) for testing are classified as “Negative”, in keeping with standard clinical practice for describing reports with no relevant clinical findings.

**Figure 1:**
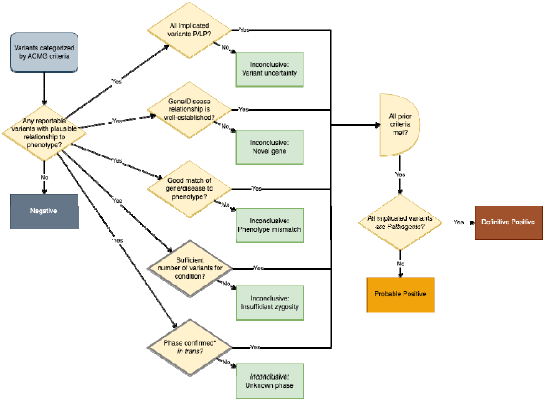
Overview of the case-level classification system. This process begins after pathogenicity categorization of individual variants. When there are no variants determined to be clinically reportable with plausible relationship to the presenting phenotype, then the case as a whole is categorized as negative. The inconclusive categories are considered in parallel: a case may be placed in the inconclusive category overall with multiple reasons applied. The inconclusive categories of insufficient zygosity or unknown phase are applied only when the condition involved has recessive mode of inheritance (dark borders in decision path). If all of the prior criteria have been met, then the categories of Probable Positive vs. Definitive Positive are applied depending on whether the implicated variant or variants are categorized as likely pathogenic or all pathogenic.

The high-level classification combines information about variant-level classification of pathogenicity; genotype, inheritance and phase; clinically-described phenotype; and gene-disease association.

### Definitive Positive

The “Definitive Positive” category is used when the combined presence of variant(s), with established phase when in combination, is sufficient to establish a diagnostic result without significant ambiguity. In all cases classified in this category, phenotype must be consistent with the condition associated with the affected genes. Operationally, this includes the identification of one Pathogenic variant in a gene that is associated with a condition with autosomal dominant inheritance. For conditions with recessive inheritance, there must be a homozygous Pathogenic variant or two Pathogenic variants with phase confirmed to be *in trans*. Cases involving X-linked or Y-linked disorders meeting this classification must have one or more Pathogenic variants with allelic state (heterozygous, hemizygous, homozygous, or compound heterozygous) that is consistent with what is known about expression of the associated condition.

### Probable Positive

The “Probable Positive” category includes cases where the preponderance of evidence indicates that the combination of genetic findings provides the explanation for the observed phenotype(s), such that the results could be considered a “Positive” result from a clinical standpoint, but there may be some small degree of uncertainty about some aspect of the genetic results. Like the “Definitive Positive” category, these cases involve good phenotypic match between the patient presentation and the known expressivity and penetrance of conditions associated with affected genes. The primary reasons for this would be cases that would otherwise meet the criteria for “Definitive Positive”, but where one or more implicated variants are classified as Likely Pathogenic rather than Pathogenic. For example, a single “Likely Pathogenic” variant in a gene with autosomal dominant inheritance, or combination of two Likely Pathogenic or one each of Pathogenic and Likely Pathogenic variants, with demonstrated phase *in trans* in an autosomal recessive condition.

The SADY working group also considered exceptional cases where there were a combination of two Pathogenic or Likely Pathogenic variants in a gene with good phenotypic concordance, but in which phase could not be definitively determined because of the inability to test multiple informative family members. This can occur when only one parent or sibling can be tested, but is found to have one of the implicated variants. It is unlikely but possible in such situations that the second variant is *de novo* and *in cis* on the same allele as the variant that was determined to be familial, but it is much more likely that the variants are *in trans*, so this result is considered Probable Positive for the purposes of informing clinical management. As a local modification to the metric, one site reported cases as Probable Positive when a Pathogenic or Likely Pathogenic variant was detected *in trans* with a Variant of Uncertain Significance and there were no other plausible findings.^18^

### Inconclusive

The individual factors leading to a case being classified as Inconclusive are not mutually exclusive. An individual case may have ambiguity in the overall result from some combination of variant-level uncertainty, insufficient zygosity, phenotype mismatch, implication of a novel gene-disease relationship, or inadequate determination of phase.

#### Variant-level uncertainty

can contribute to a case being reported as inconclusive when a single Variant of Uncertain Significance (VUS) is identified in a gene associated with a dominant condition under consideration or when one or both variants identified in a gene associated with a recessive condition are classified at the variant-level as VUS. The decision as to whether to include VUS in a report is based on the laboratorian’s assessment of other factors, such as potential relatedness to the described phenotype, or the potential to adjudicate the variant by additional clinical evaluation.

#### Insufficient zygosity

is implicated when the reported variant, on its own, would not be sufficient to account for the phenotype based on existing knowledge of the mode of inheritance of the associated condition. The laboratory may report these variants considering the possibility that the performed testing may not have been sensitive enough to identify another causative variant in the same gene.

#### Phenotype mismatch

can refer either to expansion or contraction of the known phenotype associated with a condition in comparison to the described patient’s phenotype. Because the CSER2 studies differed with respect to granularity of phenotype data available to laboratories and procedures for evaluating patients for the different features of a condition, apparent “phenotypic contraction” may result from a lack of knowledge, at the time of laboratory interpretation, of features that are currently expressed in the patient, or could reflect features that are not yet evident because of age-related penetrance. Conversely, phenotype mismatch can also be used to indicate that there are aspects of the patient’s phenotype that cannot be explained by what is currently known about the gene-disease association. Classifying such cases as inconclusive could prompt the clinician to consider whether the unexplained features are unrelated (e.g., possibly due to another condition), or are a rare manifestation of the reported condition that has not yet been well-described.

Laboratories may choose to report some findings in which the gene-disease relationship has not been definitively established (implicating a potential **novel gene** to disease relationship). Inclusion of such findings should involve substantial preliminary evidence, such as the burden of suspected-deleterious variants in other individuals with similar clinical features, model organism data, or pathway modeling. Given the tentative nature of the undefined gene-disease associations, such cases are classified as inconclusive.

When phase cannot be adequately determined to establish that there is biallelic effect in a gene associated with a recessive condition (e.g. no parent available for testing), cases are classified as inconclusive because of **unknown phase**. Laboratory reports in these cases may recommend additional studies for phasing, such as long-range sequencing, allele-specific sequencing, or targeted testing of family members which may resolve the uncertainty of these cases.

#### Negative

Negative cases are those for which there are no findings that are considered positive nor which rose to the level of plausibility for reporting as inconclusive. This classification is provided for the classification of the genetic findings with respect to the phenotypic indication(s) for which testing was performed. For instance, an individual with only secondary findings would have testing classified as negative for the primary diagnostic analysis. As with any diagnostic test, a classification of “negative” does not necessarily rule out a genetic condition. Since this classification is for the laboratory itself, the assessment of post-test probability that there could still be an underlying genetic condition would include clinical assessment of prior probability of a genetic condition given the phenotypes observed, and the expected sensitivity and specificity of genome-scale sequencing for evaluating conditions with those phenotypes.

### Use of case-level results categories across CSER2 sites

A total of 3389 participant cases were reported in the final May 2023 consortium-level data freeze (Table 2). For sites that recorded multiple classifications over time, the most recent classification at the time of that data freeze was used. For sites that recorded case-level classifications for multiple indications for testing, data are aggregated to the most clinically significant of the major categories (Definitive Positive > Probable Positive > Inconclusive > Negative). Clinical diagnostic yield was calculated across the consortium using the combination of cases classified Definitive Positive (n=345) and Probable Positive (n=170), for an overall diagnostic yield of 15.2% (Table 2). Yield of reportable results (cases with at least one reportable finding, Definitive Positive, Probable Positive, or Inconclusive) was 45.1% across the consortium.

**Table 2:** Summary of cases by site and overall classification. The number of cases reported at each site are subdivided into those considered clinically diagnostic (combination of Definitive Positive and Probable Positive categories), Inconclusive (with any combination of subcategories), or Negative (those with no reportable results). Diagnostic yield represents the percentage of cases where a clinically diagnostic result was reported. Reportable yield provides the percentage of cases where a diagnostic or inconclusive result was returned. The upper-right triangle provides p-values for pairwise comparisons of diagnostic vs. non-diagnostic findings (two-sided Fisher’s exact test with Bonferroni correction), and the lower-left provides p-values pairwise comparisons of use of the three aggregate categories (chi-squared tests with Bonferroni correction). Bold is used to show p-values less than 0.05. All calculations were performed using SciPy 1.16.3.^23^

| Site | Number of cases |  |  | Yield of findings |  | Pairwise comparison of classifications (p-values) |  |  |  |  |  |
| --- | --- | --- | --- | --- | --- | --- | --- | --- | --- | --- | --- |
|  | Positive | Inconclusive | Negative | Diagnostic | Reportable | CHARM | KidsCanSeq | NCGENES 2 | NYCKidSeq | P3EGS | SouthSeq |
| CHARM | 27 | 68 | 732 | 3.3% | 11.5% |  | 5.1E-04 | 1.0E+00 | 1.0E-22 | 6.0E-27 | 1.7E-43 |
| KidsCanSeq | 49 | 388 | 139 | 8.5% | 75.9% | 7.6E-132 |  | 1.0E+00 | 3.1E-06 | 2.6E-09 | 2.3E-18 |
| NCGENES 2 | 7 | 48 | 44 | 7.1% | 55.6% | 1.3E-29 | 2.1E-03 |  | 6.0E-02 | 4.4E-03 | 1.3E-05 |
| NYCKidSeq | 133 | 366 | 218 | 18.5% | 69.6% | 1.6E-118 | 1.1E-08 | 3.5E-02 |  | 1.0E+00 | 3.0E-04 |
| P3EGS | 118 | 54 | 360 | 22.2% | 32.3% | 3.2E-27 | 1.5E-81 | 1.2E-19 | 7.2E-53 |  | 2.8E-01 |
| SouthSeq | 181 | 88 | 369 | 28.4% | 42.2% | 2.4E-46 | 3.4E-79 | 1.4E-15 | 1.1E-45 | 3.7E-02 |  |
| Total | 515 | 1012 | 1862 | 15.2% | 45.1% |  |  |  |  |  |  |

Overall rates of clinically diagnostic cases had statistically significant differences across sites (chi-squared test; *p*=2.0×10^-7^). Many sites also showed statistically significant differences in clinical diagnostic yield with other sites with pairwise comparison using two-sided Fisher’s exact test. Differences in yield among NYCKidSeq, P^3^EGS, and SouthSeq were not statistically significant, nor between NCGENES 2 and KidsCanSeq or CHARM (Table 2, upper right triangle of pairwise comparisons). Pairwise comparison between the diagnostic yield of CHARM and KidsCanSeq was statistically significant at *p*=5.1×10^-4^.

Among the 1012 cases (29.9%) classified as having reportable inconclusive findings, 883 (26.1%) were described with one inconclusive subcategory, 114 (3.4%) with two subcategories, and 15 (0.4%) with three subcategories (Figure 2). There were no cases with reportable inconclusive results that included more than three subcategories. The most common subcategory used to describe inconclusive results was variant uncertainty, applied to a total of 916 cases (27.0%), and being the sole subcategory applied in 805 cases (23.8%).

**Figure 2:**
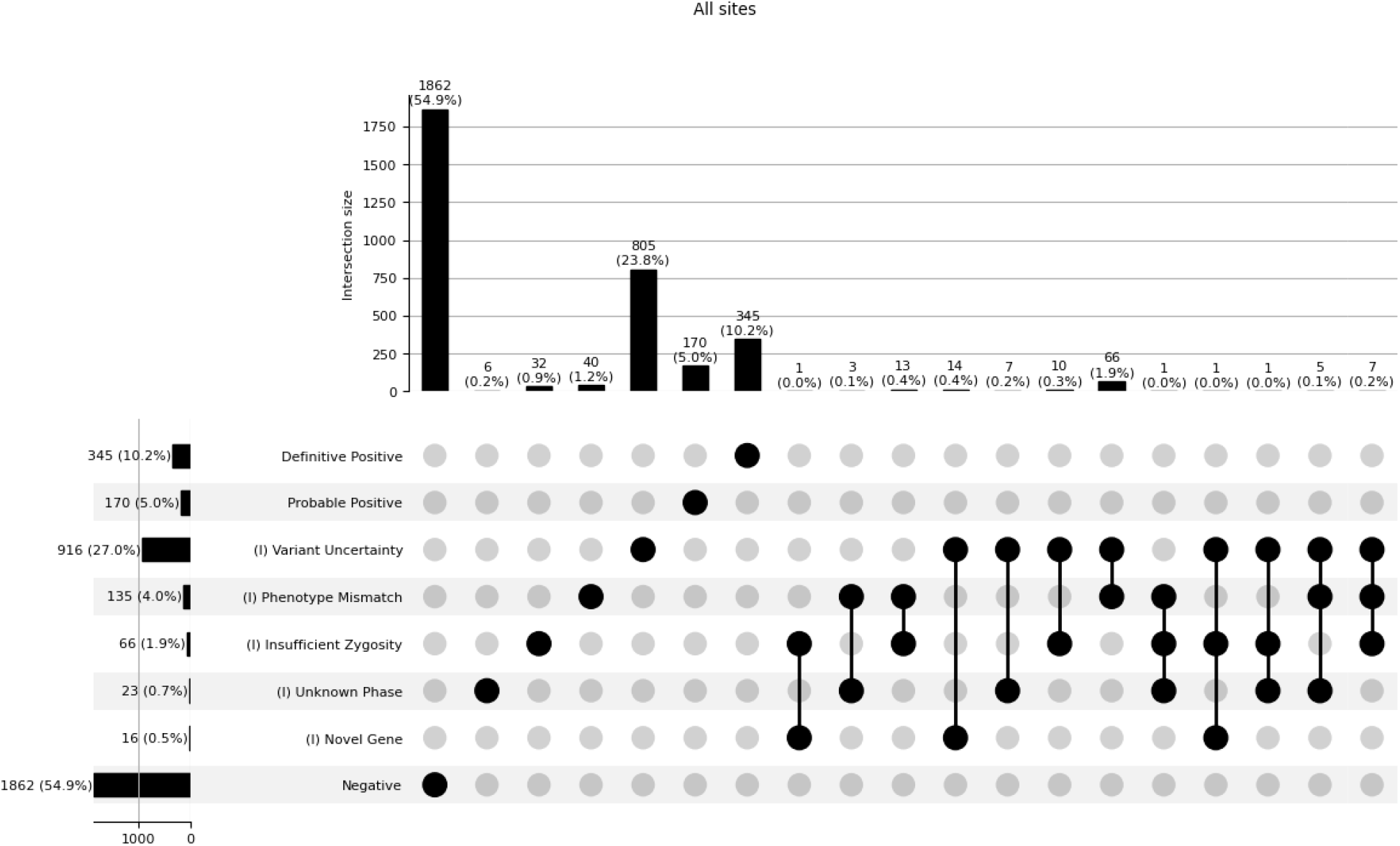
Categorized case-level results across CSER2 consortium. Data are summarized as UpSet plots showing case counts with specific classifications (in rows) and with combinations of classifications (columns). Similar figures for categorization of results for individual sites are provided in Supplemental File 1.

## Discussion

The interpretation of whether a collection of identified genetic variants explains one or more aspects of a patient’s clinical presentation incorporates factors such as certainty about variant pathogenicity, the combination of variants identified (and their phase), the gene-disease relationship, and correlation to the patient’s phenotype. At the level of clinical practice, it is this interpretation, as initially made by the reporting laboratory and then contextualized by the ordering provider, that will guide future medical management. Enabling better characterization of the way these assessments are made and the implications of this process on later clinical management was a central goal of the CSER2 consortium.

As is the case for reporting of individual variants, there is no clear boundary for the threshold between which results are considered clinically reportable and not clinically reportable. Laboratory practices differ with regard to which variants of uncertain significance may be reported to clinicians for return to individuals undergoing genetic testing. In the case of targeted panel testing, in which sequencing is being performed in response to a known relationship of a relatively small set of genes to the presenting phenotype, it is reasonable to report all variants that are classified as VUS. However, when exome or genome sequencing studies are performed, the vast majority of analytically-detectable VUS have no expected relevance to the indication for testing, so variants may be prioritized by expected contributions to the patient’s clinical characteristics and the potential usefulness of reported VUS for later analysis. With regard to VUS, some laboratories subclassify through combined evidence to determine likelihood of reclassification to pathogenic or likely pathogenic. This clinical decision-making at the level of reporting also holds for other factors that might lead a result to be inconclusive. For example, single pathogenic variants in genes associated with recessive conditions with good correlation to the patient’s presentation may be reported, accounting for the possibility of an undetected or currently unclassified variant on the other allele that may be identified in subsequent analysis.^24^ However, carrier status for conditions unrelated to the presentation are not generally recommended for report.^25^

The use of this case-level metric not only provides description of the diagnostic yield across the different sites in the consortium (as measured by proportion of cases with definitive positive or probable positive results), but also reveals other differences in the reporting of inconclusive results that were consistent with qualitative differences in site study design or study populations (Table 2). While lack of complete concordance even with respect to the classification of variant results is well-described^3^, we mitigated potential differences in classification at the variant and case level through shared discussion of cases that were challenging to categorize. Because of this mitigation, while the overall comparisons of proportions of diagnostic, inconclusive, and negative results were statistically significant across all pairwise comparisons of sites, we were able to examine differences in use of the inconclusive subcategories with respect to participant characteristics and other factors of study design. The CHARM study involved assessment for hereditary cancer risk as a pilot population-level screen, and as such the prior probability of a diagnostic genetic result was low, reflected in the diagnostic yield (3.3%) and increased stringency for considering results with factors that could contribute to diagnostic uncertainty (with variant uncertainty as the only inconclusive category used in CHARM). The KidsCanSeq and NCGENES2 studies also had low prior probability of a germline genetic condition because of their study populations. KidsCanSeq enrolled children with pediatric cancer to identify rare cases of germline cancer predisposition either as a cause of these cancers or that might impact treatment and subsequent surveillance. The reported diagnostic yield related to cancer predisposition in this cohort (8.5%) is consistent with the fact that the majority of cancer, even in pediatric cases, does not have an apparent germline predisposition. KidsCanSeq also implicated phenotype mismatch in 6.2% of cases (Supplemental Document 1), since they reported variants in genes associated with cancer predisposition even if not associated with the child’s specific cancer diagnosis^13^. The NCGENES2 study enrolled pediatric participants at the first outpatient visit for a pediatric genetics or neurology visit because of suspicion of a genetic condition. The lower pre-test risk in this study due to lack of prior clinical selection for higher likelihood of a genetic condition, coupled with a relative paucity of phenotypic information (especially for conditions with age-related expression) is also consistent with the lower overall diagnostic yield (7.1%) compared to other populations in which exome sequencing has been performed, and higher implication of phenotype mismatch (18.2%; Supplemental Document 1) compared to the other studies in the consortium. The NYCKidSeq, P^3^EGS and SouthSeq studies had mixtures of patient populations that more closely reflected current clinical indications for genome-scale sequencing, with diagnostic yield similar among these sites (18.5%, 22.1%, 28.4%, respectively).

Across all sites, variant uncertainty was the largest contributor of inconclusive results, either individually or in combination with other factors (27.0% of overall cases in the consortium, ranging from 8.2% in CHARM to 59.5% in KidsCanSeq). The difference in threshold for determining which VUS are considered “reportable” also contributed to the variability in reportable yield (Table 2), again paralleling study design decisions regarding the potential that the communication of a VUS might inform diagnostic thinking or enable additional clarifying evaluation. Phenotype mismatch was the second most common factor described in inconclusive results (4.0% of overall cases), but also showed substantial variability, with highest rate in NCGENES 2 (18.2%) consistent with the fact that participants early in the course of their diagnostic odysseys may not have developed clinical signs or not yet had studies (e.g., radiographic imaging) to show findings to more clearly fit or rule out the expected phenotype for a condition. Although there was no set threshold for number of combined inconclusive factors reported, it was rare for sites to report cases with 3 inconclusive categories (0.5% in KidsCanSeq, 6.0% in NCGENES2, 0.6% in NYCKidSeq, 0.4% in PEGS, none in other sites), and no sites reported cases with more than three inconclusive categories, demonstrating an empiric limit to the number of factors that laboratories might be willing to accept in a reportable result. Unknown phase was also a rarely reported category, but might occur when parental samples were not available to verify phase and in the absence of long read sequencing data.

One potential benefit of reporting inconclusive results is the potential to guide further clinical evaluation or other molecular studies to clarify the results. The probability of clinical follow-up is difficult to predict in the context of a specific result, but we expect that follow-up may vary depending on the reasons given for an inconclusive result. Results classified as inconclusive due to unknown phase may be more easily resolvable than others if family members are available for genetic testing. Reporting of results that are inconclusive due to phenotypic mismatch may lead to additional clinical evaluation to look for the presence of features that were not reported to the lab at the time the results were classified. Results that are inconclusive due to uncertainty at the level of the gene-disease association would only be resolved with additional information about that association, such as through accumulation of additional cases.

The ability to quantify the contributions of different factors to case-level ambiguity through categories of inconclusive results is an important step in improving understanding the implications of this ambiguity on the clinical usefulness of downstream results. In addition to the use of this metric in the site-level reporting under CSER2 (Table 1) and aggregate analysis of factors affecting diagnostic yield among the sites performing diagnostic testing (manuscript submitted), it has been adopted by a consortium studying the impact of prenatal genome sequencing (PrenatalSeq) and by a research service core facility (GENYSIS) that provides clinical analysis of genome sequence data obtained through research studies^26^. We expect that this rubric for categorization will be useful to the broader genetics community in both clinical and research settings to standardize the case-level representation of genetic test results, providing a level of meaning that expands beyond variant-level results.

## Data Availability

This CSER2 standardized dataset was aggregated by the consortium data coordinating center and is available through the NHGRI Analysis Visualization and Informatics Lab-space (AnVIL; https://anvilproject.org/consortia/cser/resources). Access to CSER2 data is moderated through dbGaP data access requests. The following phsIDs are associated with CSER consortium data: phs001089. v3.p1 (SouthSeq), phs002110.v1.p1 (NCGENES 2), phs002111.v1.p1(CHARM), phs002324.v1.p1 (P3 EGS), phs002337.v1.p1 (NYCKidSeq), phs002378.v1.p1 (KidsCanSeq).

