## Supplemental Figures for "Measuring the Meaning of Genomic Results: Harmonization of the Metric for Case-Level Results in the CSER2 Consortium"

**Supplemental document:** Per-site UpSet plots of case results by granular categories, including subdivision of inconclusive results.

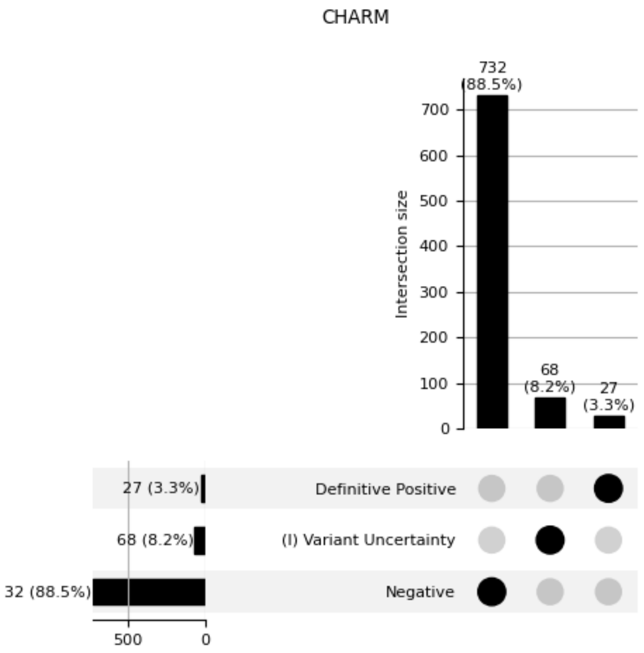

KidsCanSeq

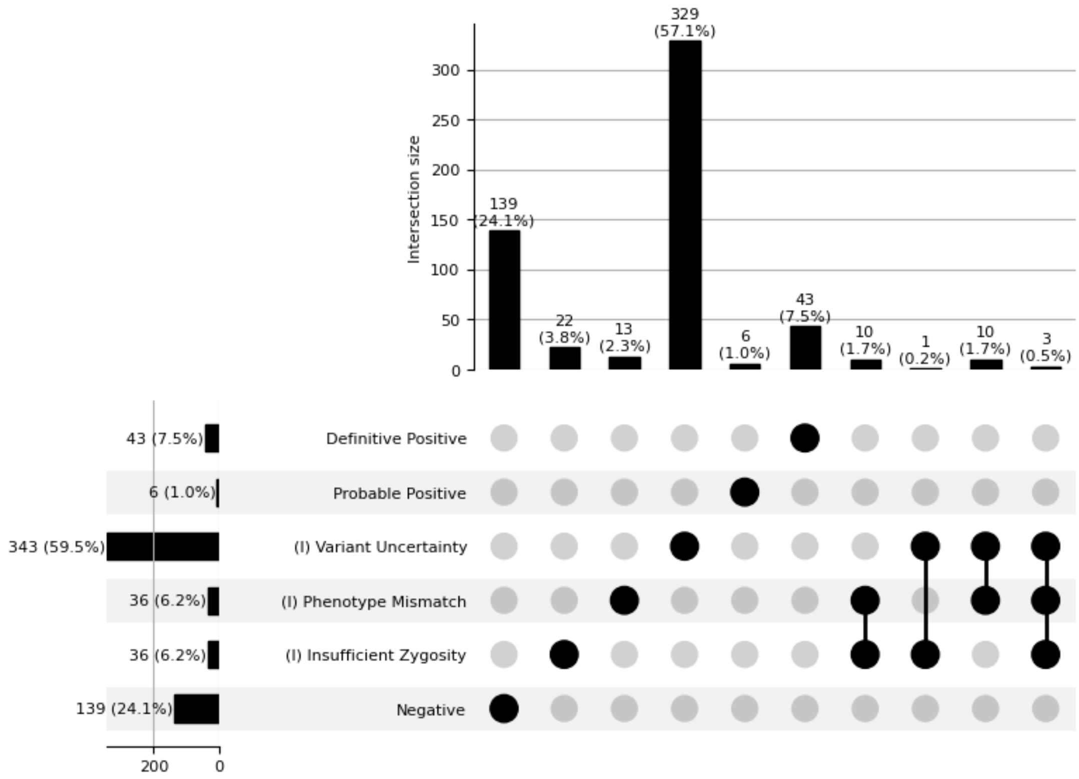
Intersection size
0
50
100
150
200
250
300
139 (24.1%)
22 (3.8%)
13 (2.3%)
329 (57.1%)
6 (1.0%)
43 (7.5%)
10 (1.7%)
1 (0.2%)
10 (1.7%)
3 (0.5%)
43 (7.5%)
6 (1.0%)
343 (59.5%)
36 (6.2%)
36 (6.2%)
139 (24.1%)
Definitive Positive
Probable Positive
(I) Variant Uncertainty
(I) Phenotype Mismatch
(I) Insufficient Zygosity
Negative
200
0

NCGENES 2

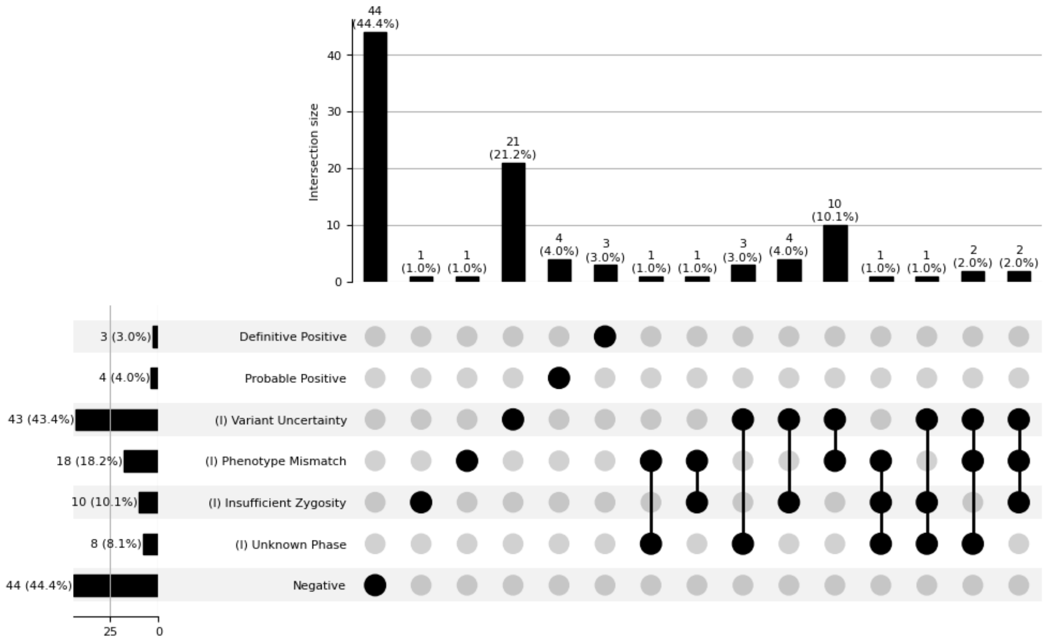


NYCKidSeq

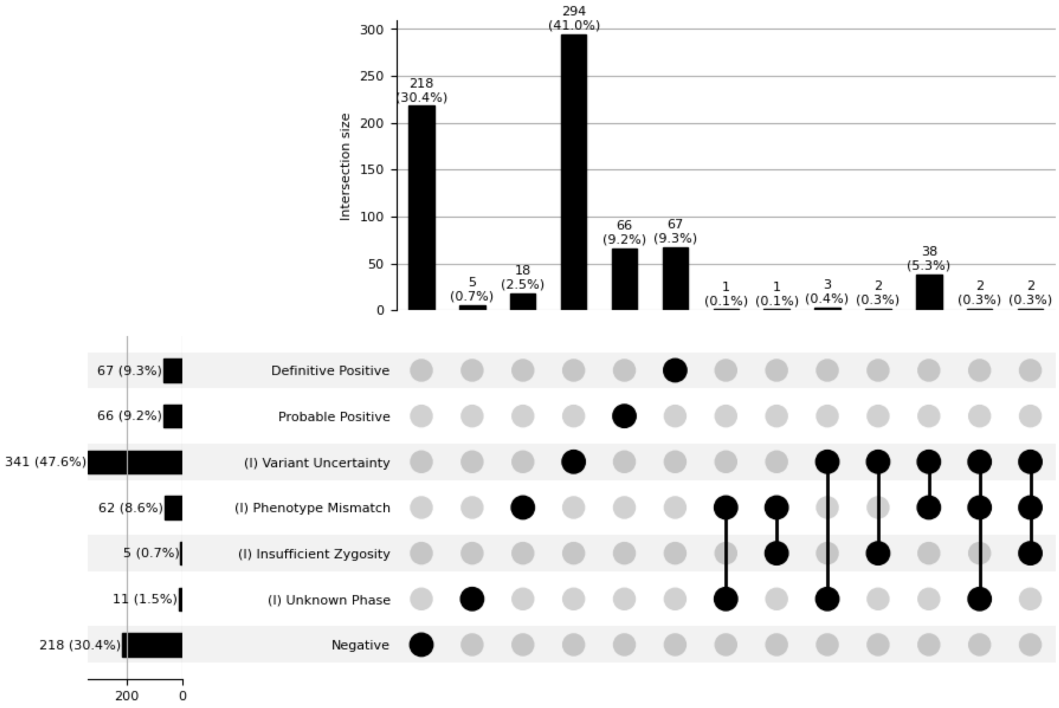

P3EGS

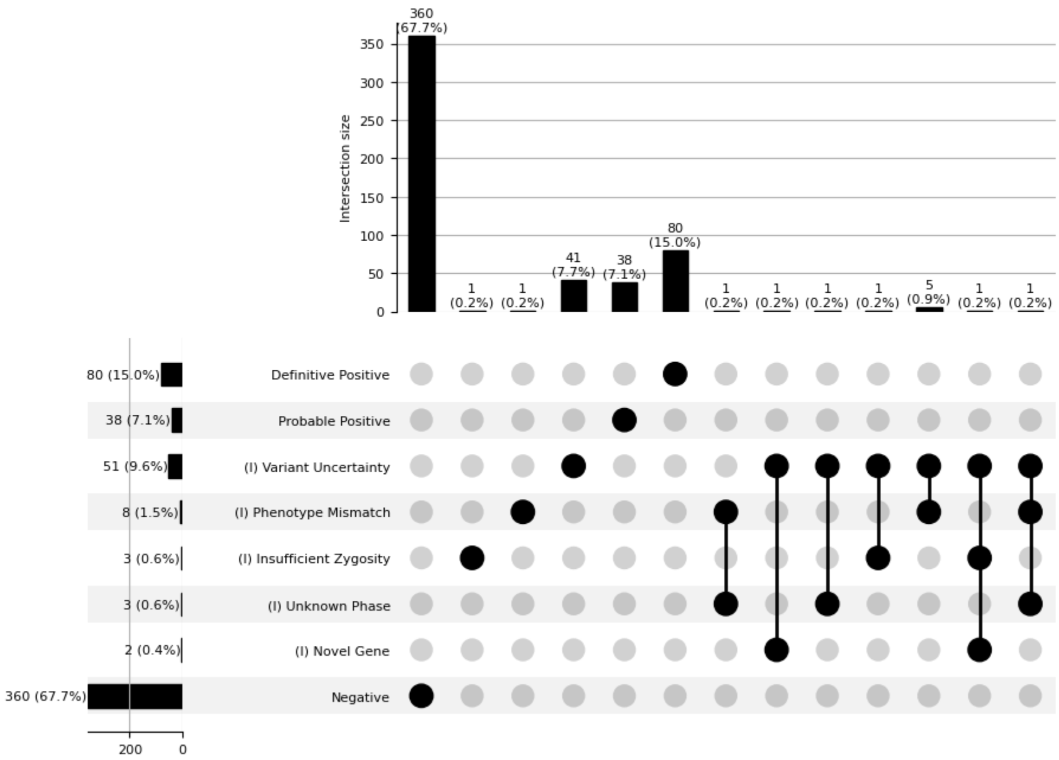

360 (67.7%)
1 (0.2%)
1 (0.2%)
41 (7.7%)
38 (7.1%)
80 (15.0%)
1 (0.2%)
1 (0.2%)
1 (0.2%)
1 (0.2%)
5 (0.9%)
1 (0.2%)
1 (0.2%)
Intersection size
0
50
100
150
200
250
300
350
80 (15.0%)
38 (7.1%)
51 (9.6%)
8 (1.5%)
3 (0.6%)
3 (0.6%)
2 (0.4%)
360 (67.7%)
Definitive Positive
Probable Positive
(I) Variant Uncertainty
(I) Phenotype Mismatch
(I) Insufficient Zygosity
(I) Unknown Phase
(I) Novel Gene
Negative
200
0

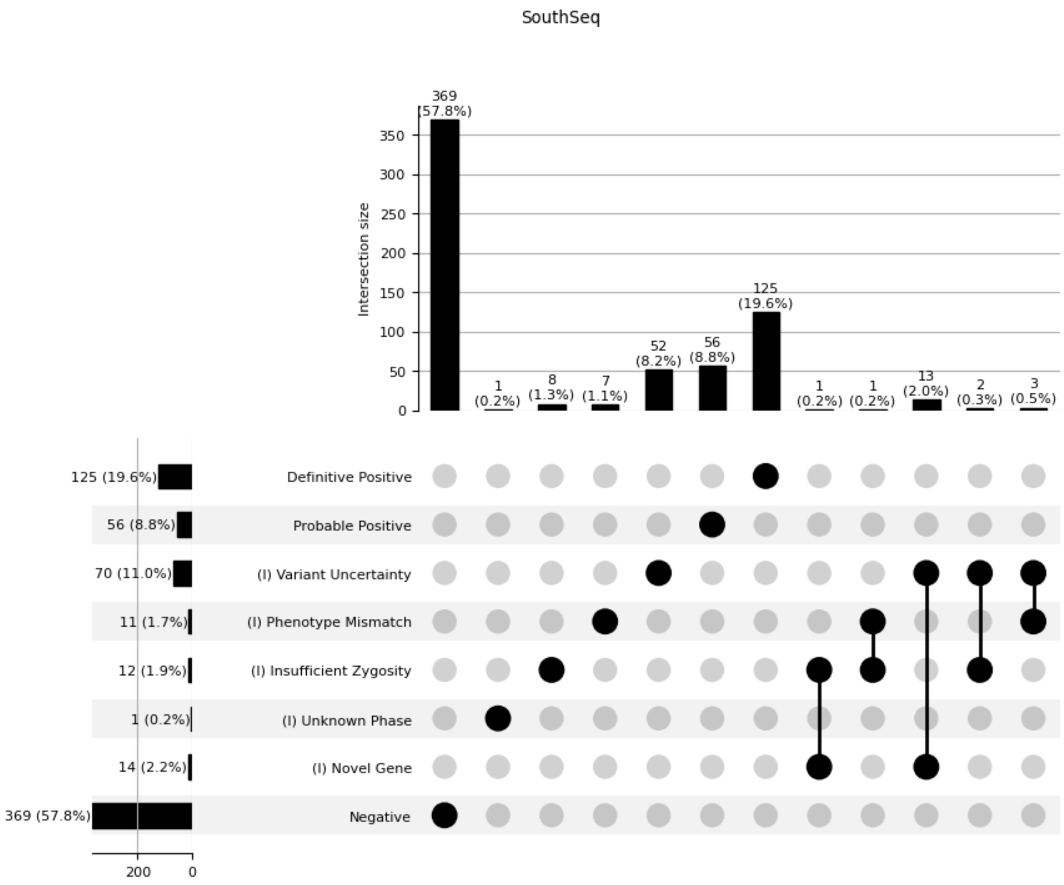
SouthSeq
Intersection size
369 (57.8%)
1 (0.2%)
8 (1.3%)
7 (1.1%)
52 (8.2%)
56 (8.8%)
125 (19.6%)
1 (0.2%)
1 (0.2%)
13 (2.0%)
2 (0.3%)
3 (0.5%)
125 (19.6%) Definitive Positive
56 (8.8%) Probable Positive
70 (11.0%) (I) Variant Uncertainty
11 (1.7%) (I) Phenotype Mismatch
12 (1.9%) (I) Insufficient Zygosity
1 (0.2%) (I) Unknown Phase
14 (2.2%) (I) Novel Gene
369 (57.8%) Negative
